# Views and experiences of young people on using mHealth platforms for sexual and reproductive health services in rural low- and middle-income countries: a qualitative systematic review

**DOI:** 10.1101/2023.09.04.23295035

**Authors:** Alexander S. Laar, Melissa L. Harris, Md N Khan, Deborah Loxton

**Affiliations:** The University of Newcastle, Australia, School of Public Health and Medicine, Centre for Women’s Health Research, Faculty of Health and Medicine, Hunter Medical Research Institute, Callaghan, New South Wales 2308, Australia.

**Keywords:** Healthcare professionals, young people, mHealth platforms, rural settings, LMICs

## Abstract

In low- and middle-income countries (LMICs), reproductive health programs use mobile health (mHealth) platforms to deliver a broad range of SRH information and services to young people in rural areas. However, young people’s experiences of using mobile phone platforms for SRH services in the rural contexts of LMICs remains unexplored. This review qualitatively explored the experiences and perceptions of young people’s use of mobile phone platforms for SRH information and services. This qualitative evidence synthesis was conducted through a systematic search of online databases: Medline, Embase, CINAHL, PsycInfo and Scopus. We included peer reviewed articles that were conducted between 2000 and 2020 and used qualitative methods. The methodological quality of papers was assessed by two authors using Grading of Recommendations, Assessment, Development and Evaluation (GRADE) and Confidence in Evidence from Reviews of Qualitative research (CERQual) approach with the identified papers synthesized using a narrative thematic analysis approach. The 26 studies included in the review were conducted in a wide range of LMIC rural settings. The studies used seven different types of mHealth platforms in providing access to SRH information and services on contraception, family planning, sexually transmitted infections (STIs) and human immunodeficiency virus (HIV) education. Participant preferences for use of SRH service platforms centred on convenience, privacy and confidentiality, as well as ease and affordability. High confidence was found in the studies preferencing text messaging, voice messaging, and interactive voice response services while moderate confidence was found in studies focused on phone calls. The overall constraint for platforms services included poor and limited network and electricity connectivity (high confidence in the study findings), limited access to mobile phones and mobile credit due to cost, influence from socio-cultural norms and beliefs and community members (moderate confidence in the study findings), language and literacy skills constraints (high confidence in the study findings). The findings provide valuable information on the preferences of mHealth platforms for accessing SRH services among young people in rural settings in LMICs and the quality of available evidence on the topic. As such, the findings have important implications for health policy makers and implementers and mHealth technology platform developers on improving services for sustainable adoption and integration in LMIC rural health systems.

## Introduction

Sexual and reproductive health (SRH) is essential to young people’s overall health and wellbeing globally (1, 2). While the 1994 Conference on Population and Development recognised the rights of young people to access SRH services, including contraception and family planning [2], in low- and middle-income countries (LMICs) most young people aged 10–24 years have very limited or no access to SRH information and services. Further, young people from rural settings in LMICs remain the most underserved population by SRH information and services (3-7).

Studies from rural LMICs have identified multiple challenges which limit young people’s access to and use of SRH information and services (5, 8-10). Some of these barriers include socio-cultural norms (11), stigma attached to sex and sexual health by communities (11), and religious organisations (11). In addition, challenges such as limited health resources and infrastructure, negative or unprofessional attitudes of health providers, long distances to health facilities, inability to afford cost of services (11) and long waiting times for services at health facilities deter or prevent young rural people from accessing services (4-6). Additionally, challenges of conventional SRH intervention services results in limited access to SRH (4, 12, 13). These barriers contribute to an unmet need for SRH information among young rural people and result in non-use or incorrect use of contraception and expose young people to vulnerabilities and risks which may lead to unintended pregnancies and STIs including HIV (2, 14, 15).

Meeting young people’s SRH information and service needs, especially in rural and remote areas, will require innovative modes of delivering youth-friendly services. Digital health programs offer opportunities to improve the provision and access to SRH of young people in LMICs (16). Using mobile phones for the delivery of SRH information and services in a rural context of LMICs could reduce or avert inequalities in the uptake of SRH information and services among young people in rural settings. To address the specific SRH needs of young people in remote rural settings (i.e. in hard-to-reach areas), novel and innovative mobile health (mHealth) platforms are now being used in LMICs to provide access to youth-friendly and youth-centred SRH information and services (17).

The World Health Organization defines mHealth as ‘the use of mobile and wireless technologies to support the achievement of health objectives’(17). Digital health technologies such as mobile phones can facilitate SRH information exchange and use among young people in rural LMICs to assist with making informed SRH decisions (17). It is suggested that these technologies are effective and affordable in improving coverage and access to SRH information and services among young rural people and are acceptable to young people (6, 17). However, despite the potential of mHealth platforms to improve provision and access to SRH information and services in rural settings in LMICs (17, 18), little evidence and information currently exists. What is available is qualitative in nature, which limits generalisability. A synthesis of this literature, however, can be undertaken to inform policy programs designed to address the SRH needs of young people in rural healthcare systems in LMICs. As such, we synthesised the current evidence to provide a comprehensive understanding of the views and experiences of young people in rural areas using mobile platforms for SRH information and services in rural and remote areas in LMICs.

## Methods

This systematic review is based on the Preferred Reporting Items for Systematic Reviews and Meta-Analyses (PRISMA) guidelines (19). The protocol was registered in the PROSPERO database on August 21, 2020 (# CRD42020199221).

### Data sources and search strategy

A systematic search of five online databases: Medline, Embase, CINAHL, PsycInfo and Scopus for peer-reviewed articles that reported on mHealth intervention studies with young men and women aged between 10-24 years (‘adolescents’ aged 10-19 years and ‘youth’ aged 15-24 years) (20). These two overlapping age groups were combined to describe ‘young people’ (21) from rural settings in LMICs. LMICs within this study were determined according to World Bank classifications (22). Studies were limited to those published in English from 2000-2020 given the global inception of evidence on mobile telemedicine (23) and increased interest in their use in LMICs over this period. In addition to these sources, reference lists of identified articles and key references of relevant systematic reviews were searched. Searches were also carried out in Google and Google Scholar databases. For database searches, medical subject headings (MeSH) were used to identify relevant articles. Key search terms and keywords that were used are shown in Table 1.

**Table 1:**
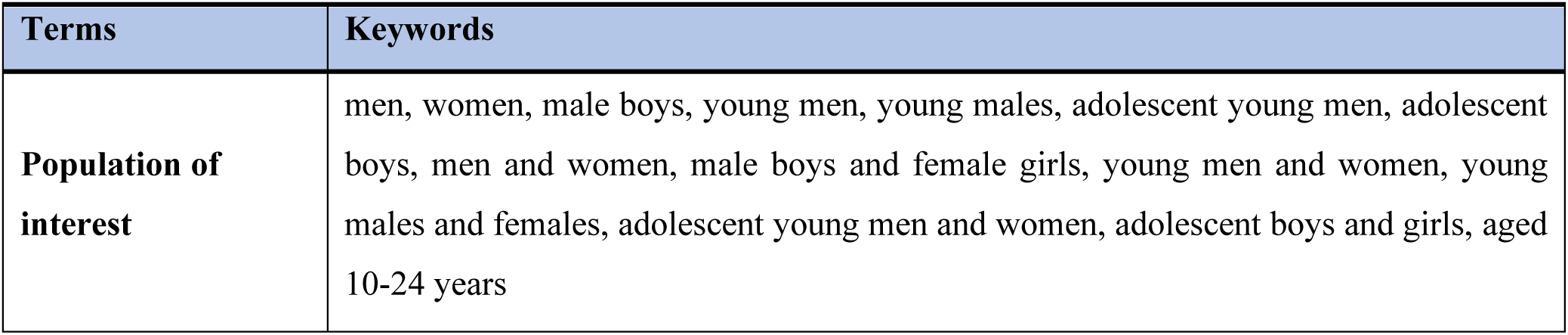
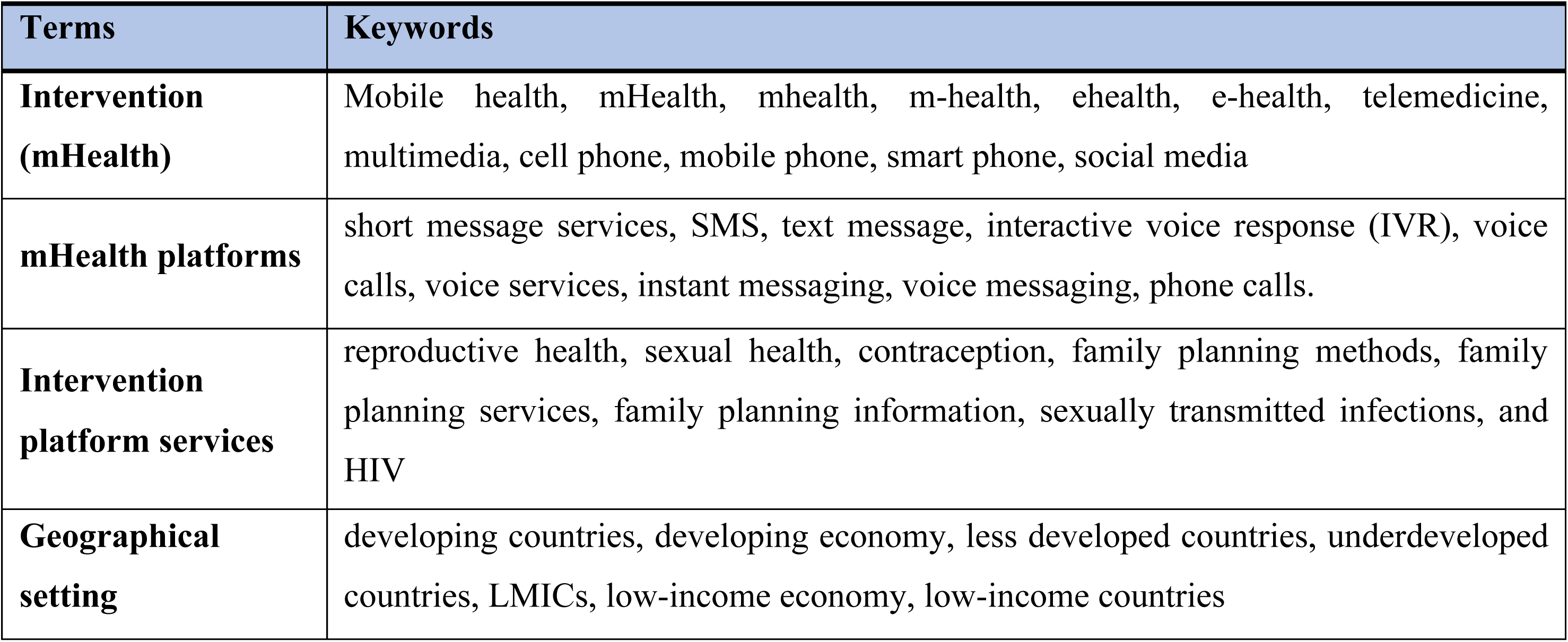
Search terms and keywords.

The first author (ASL) developed the search strategy which was reviewed by MLH and DL. The search strategy was refined in consultation with the College of Health, Medicine and Wellbeing’s librarian (see Table 1). The Medline search strategy used is shown in Table 2. Search strategies tailored to the other databases are included in the Appendix.

**Table 2:**
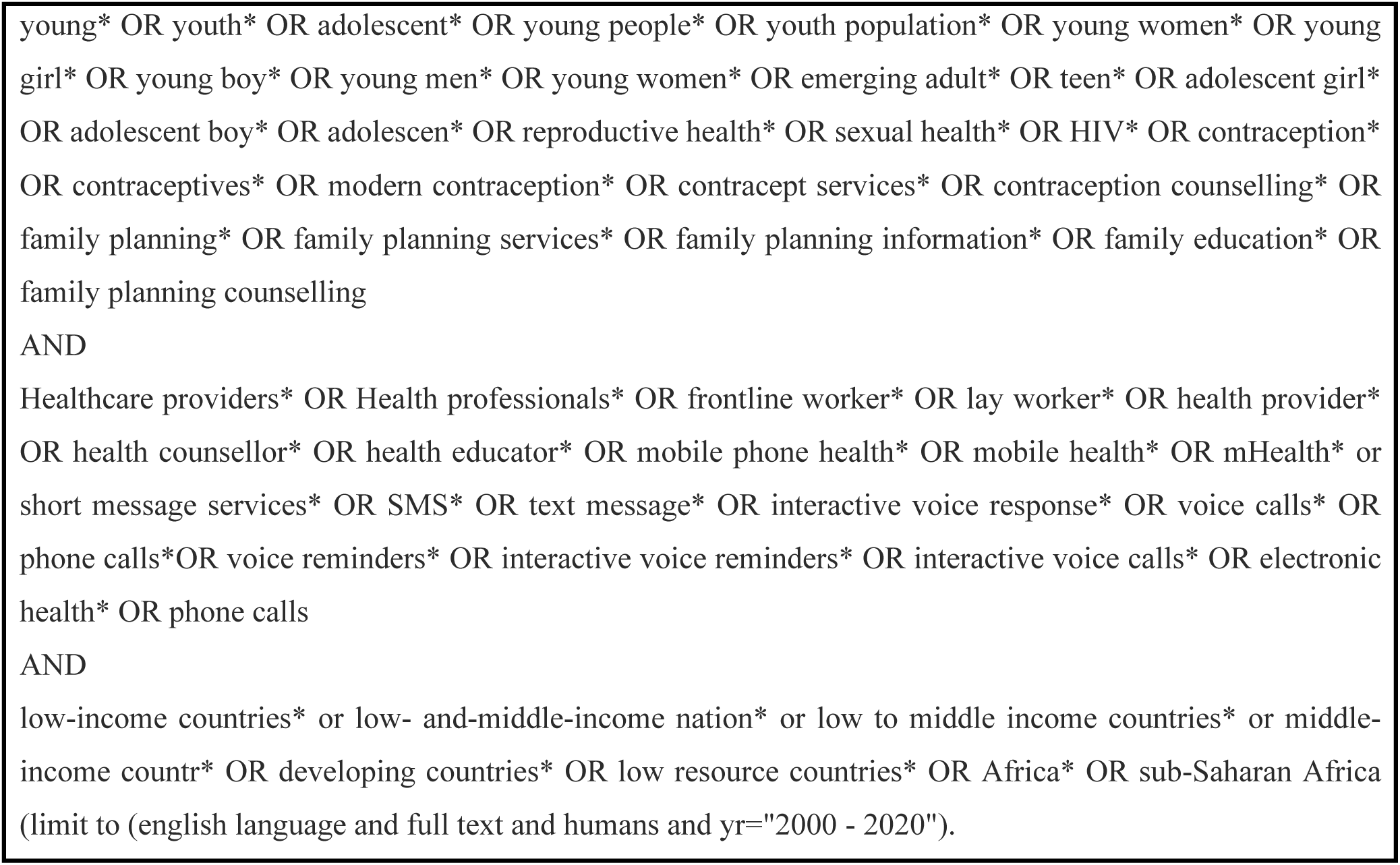
Medline search strategy.

### Inclusion and exclusion criteria

We included primary studies that used qualitative methods and reported on the views and experiences of young rural female and male’s use of mobile phone platforms for accessing SRH information and services in LMICs. For the purposes of this review, ‘young’ people are defined as (adolescents and youth populations from 10 to 24 years). Conference presentations, student theses, editorials, review articles, letters to the editor, commentaries, symposium proceedings and studies where information on young people could not be disaggregated were excluded.

### Data extraction

Database searches were completed by ASL. The literature results were first downloaded into Endnote X9 software (Thomson Reuters, Scientific Inc., New York City, New York, USA) and later imported into the Covidence online platform for analysis. Duplicates were removed and screening was then independently conducted by two reviewers (ASL and NK) based on title and abstract according to the inclusion criteria. Full text articles were retrieved and then two reviewers (ASL and NK) independently assessed the relevancy of the papers. Screened articles were read in full. Discrepancies were resolved by discussion.

### Assessment of methodological quality of studies using GRADE-CERQual approach

The methodological quality of the included studies were appraised using the Grading of Recommendations, Assessment, Development and Evaluation (GRADE) and Confidence in Evidence from Reviews of Qualitative research (CERQual) approach *(24)*. The methodological quality of included studies were appraised independently by two reviewers (ASL and NK) against four key components: methodological limitations *(25)*, coherence of the review finding *(26)*, data adequacy, *(27)* and relevance *(28)* detailed in Table 3(see also Assessment of Quality Table in Appendix). The overall CERQual explanation of quality assessment findings is also detailed in Table 4.

**Table 3:**
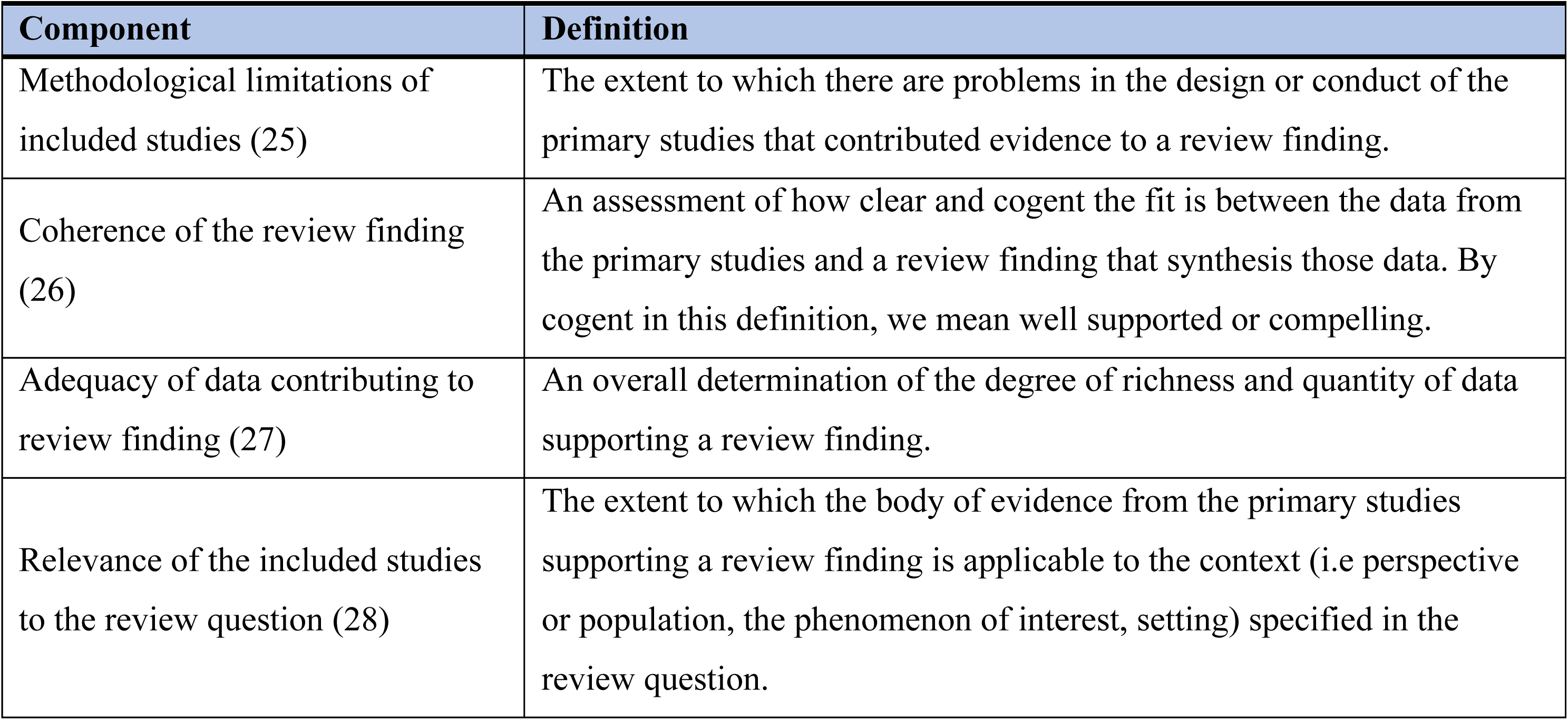
Definitions of GRADE-CERQual methodological components.

**Table 4:** Grading and rating of confidence in the individual review findings.

|  |  |
| --- | --- |
| High confidence | It is highly likely that the review finding is a reasonable representation of the phenomenon of interest. |
| Moderate confidence | It is likely that the review finding is a reasonable representation of the phenomenon of interest |
| Low confidence | It is possible that the review finding is a reasonable representation of the phenomenon of interest |
| Very low confidence | It is not clear whether the review finding is a reasonable representation of the phenomenon of interest |

### Determining the level of confidence in the study finding

We further assessed the level of rating of confidence or certainty in the study findings using the GRADE-CERQual approach in Table 4.

## Results

### Summary

A total of 3,121 articles were identified through the database search. Among these, 434 duplicates were removed. A total of 2,712 studies were assessed based on titles and abstracts. Of these, 2,672 studies were excluded for not meeting inclusion criteria. Forty full-text articles were assessed for eligibility. Of these, 14 articles were excluded. Twenty-six articles met the inclusion criteria for this review (see Figure 1).

**Figure l:**
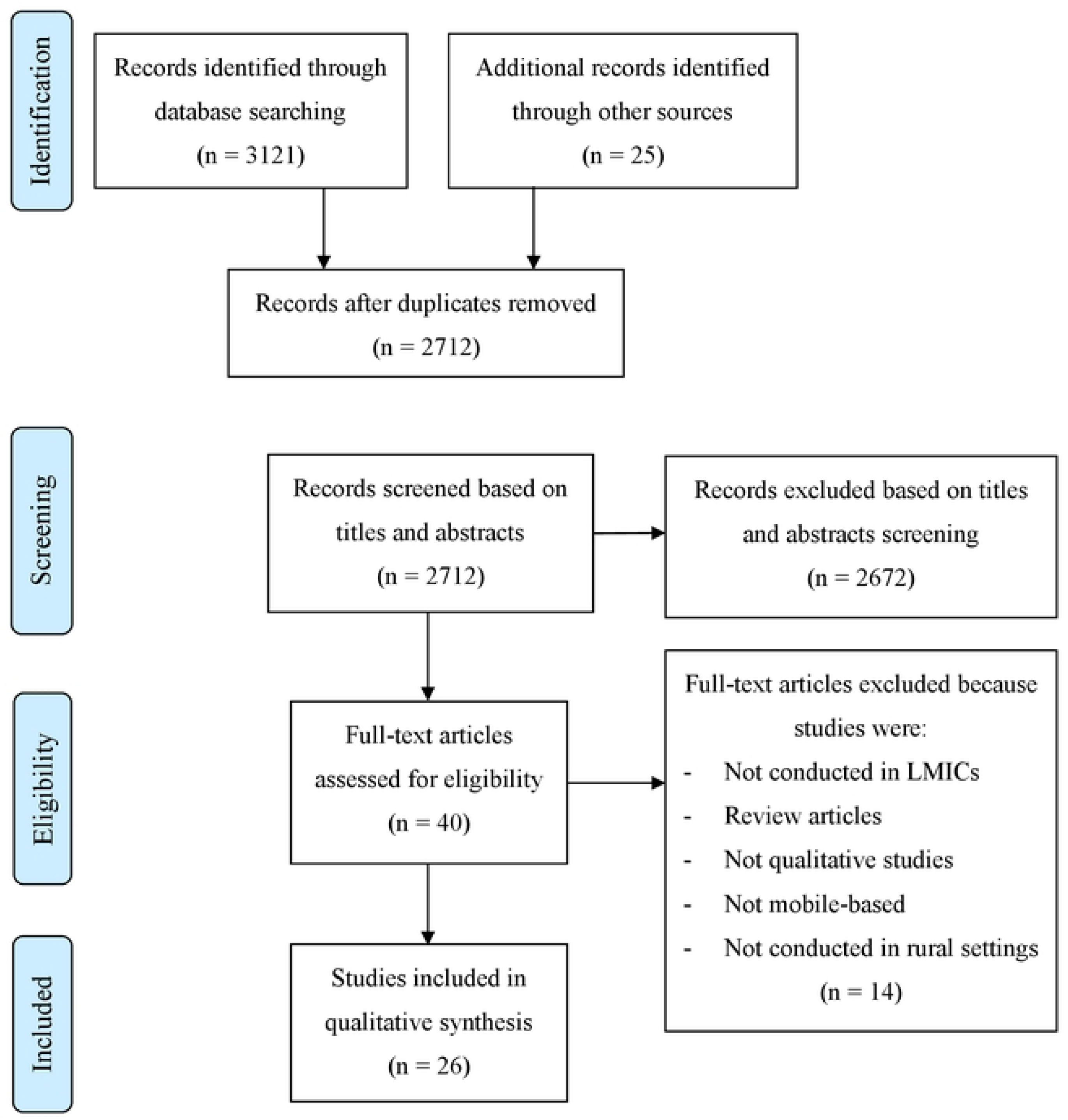
PRISMA flow chart showing study selection procedure.

### Characteristics of studies

All the included studies were conducted in rural areas in LMICs and focused on the use of mobile phone platforms for providing access to SRH information and services to young people, including contraceptive advice and information, family planning and HIV prevention education. The studies were conducted in sixteen different countries, with the majority in Kenya (8 studies) and South Africa (3 studies). Two studies were conducted in Nigeria, with an additional four in Uganda (2 studies) and Tanzania (2 studies). The remaining nine studies were conducted in Burkina-Faso, Bangladesh, Malawi (Rwanda, Malawi, and India), Cambodia, Nepal (Tajikistan, Bolivia and Palestine), Ethiopia, and Peru, respectively. In terms of the study settings, most studies recruited youth participants from communities (17 studies). Others used health care facilities (2), health care facilities and communities (2), communities and schools (3) and schools alone (2). Regarding the methods used for data collection, most involved focus groups (13 studies) and in-depth interviews (9), with four using both (see Table 5).

**Table 5:**
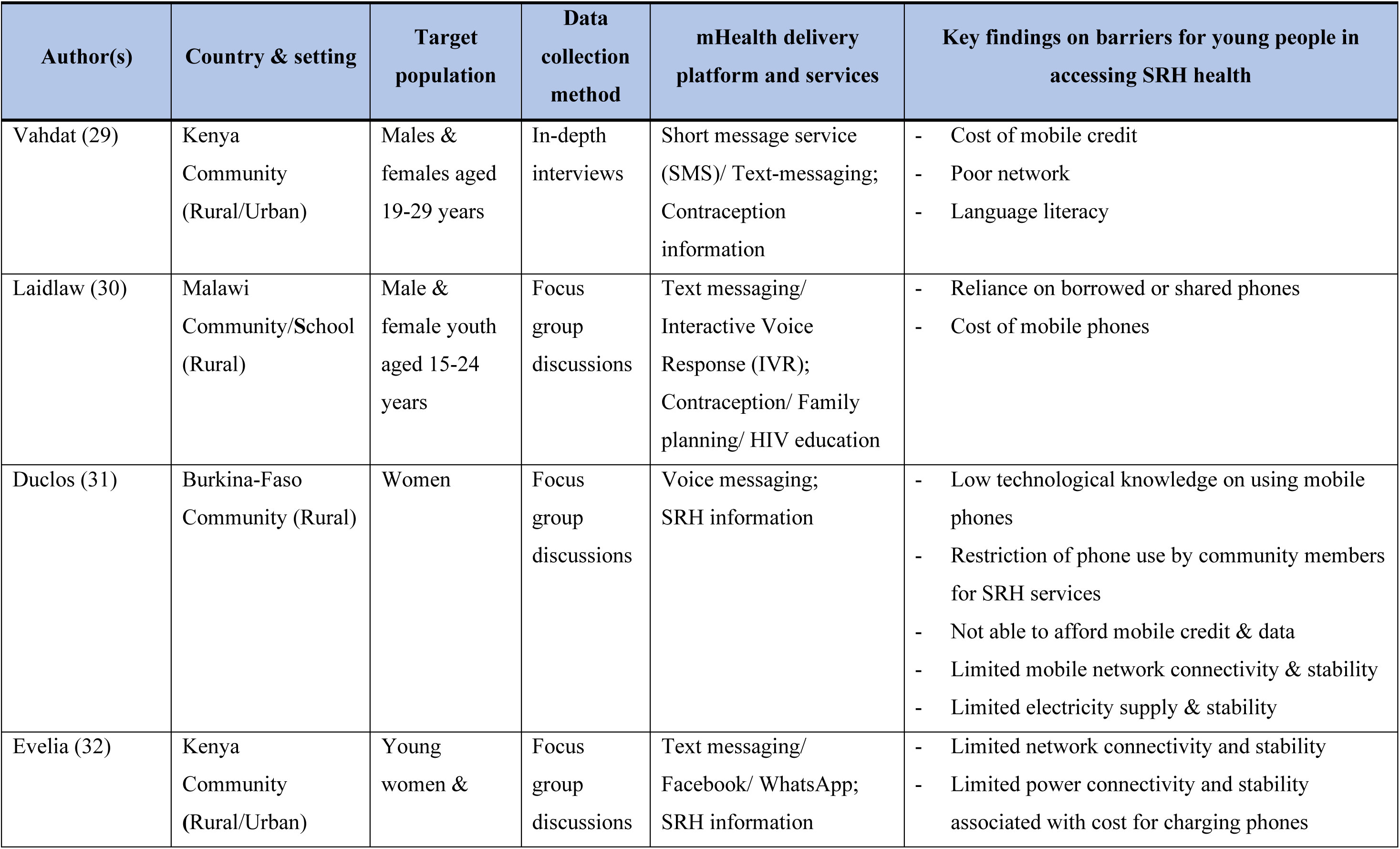
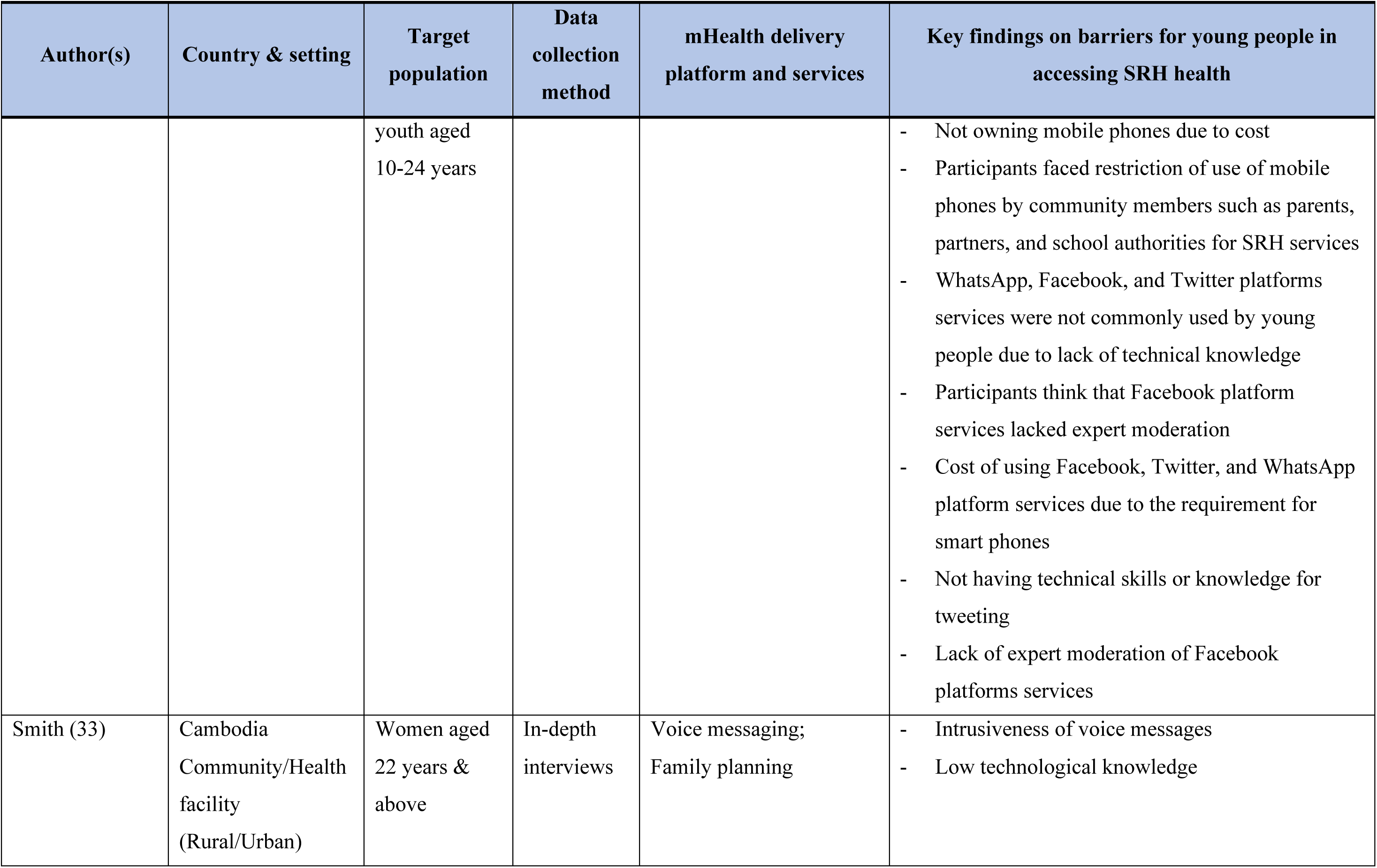
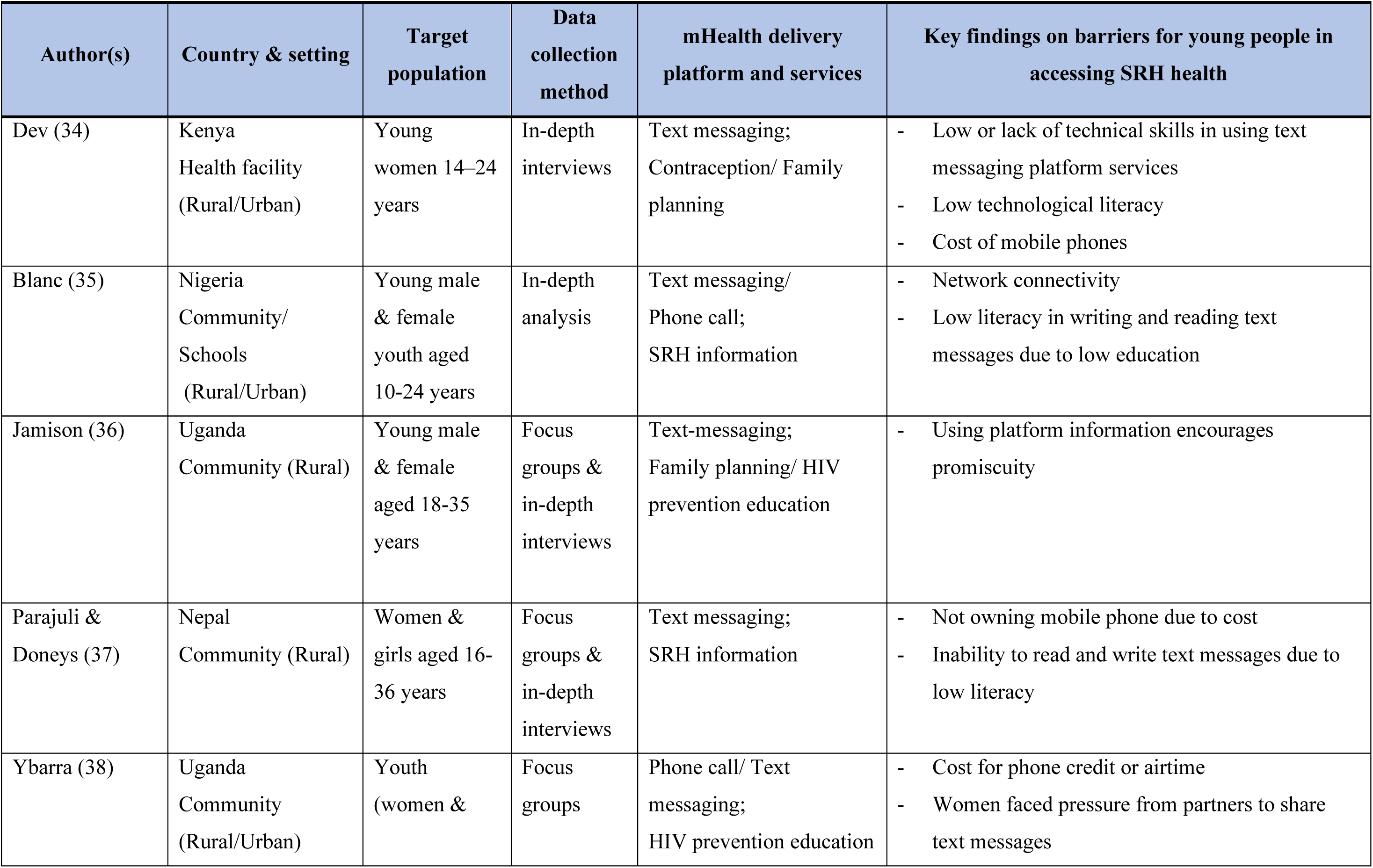
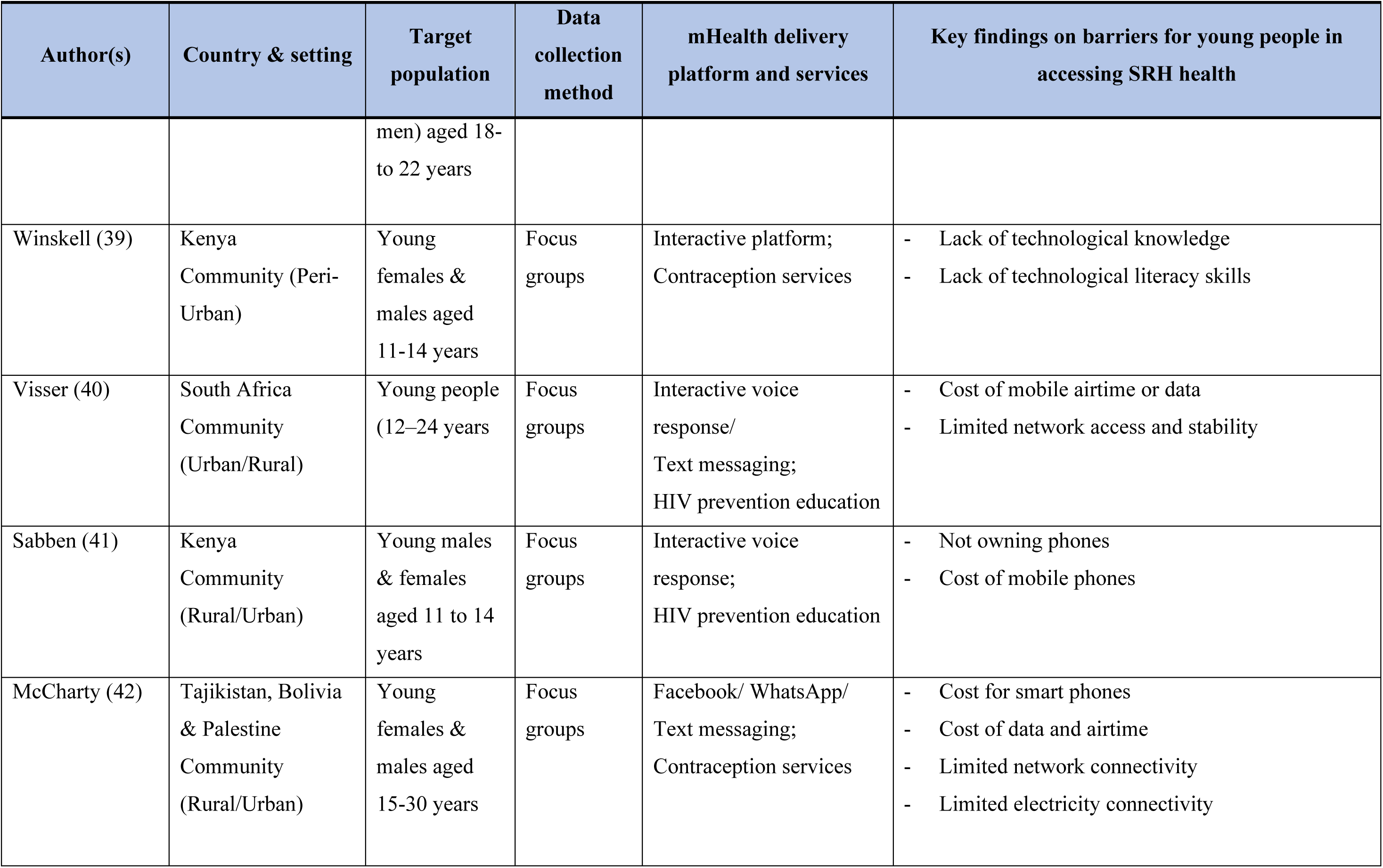
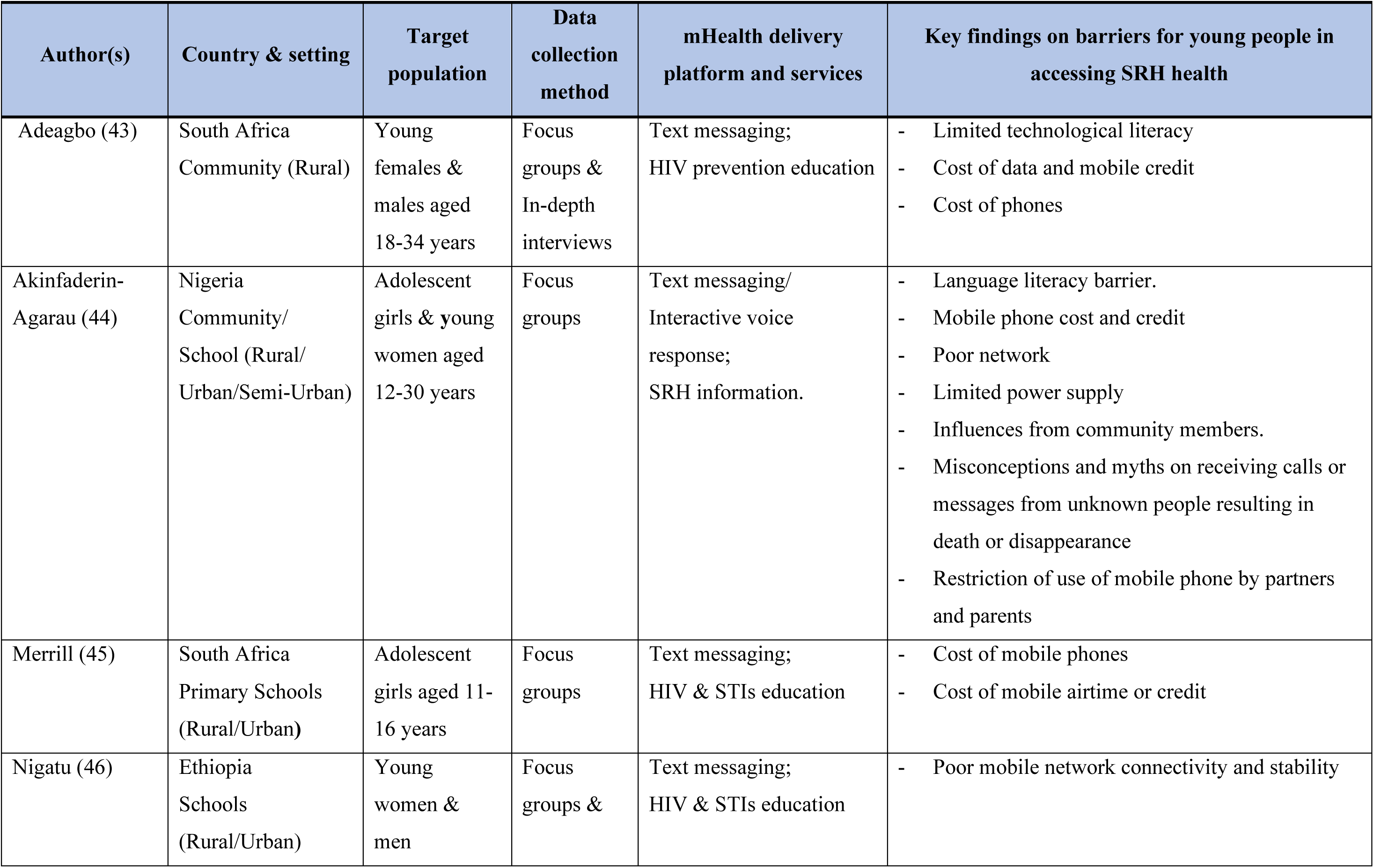
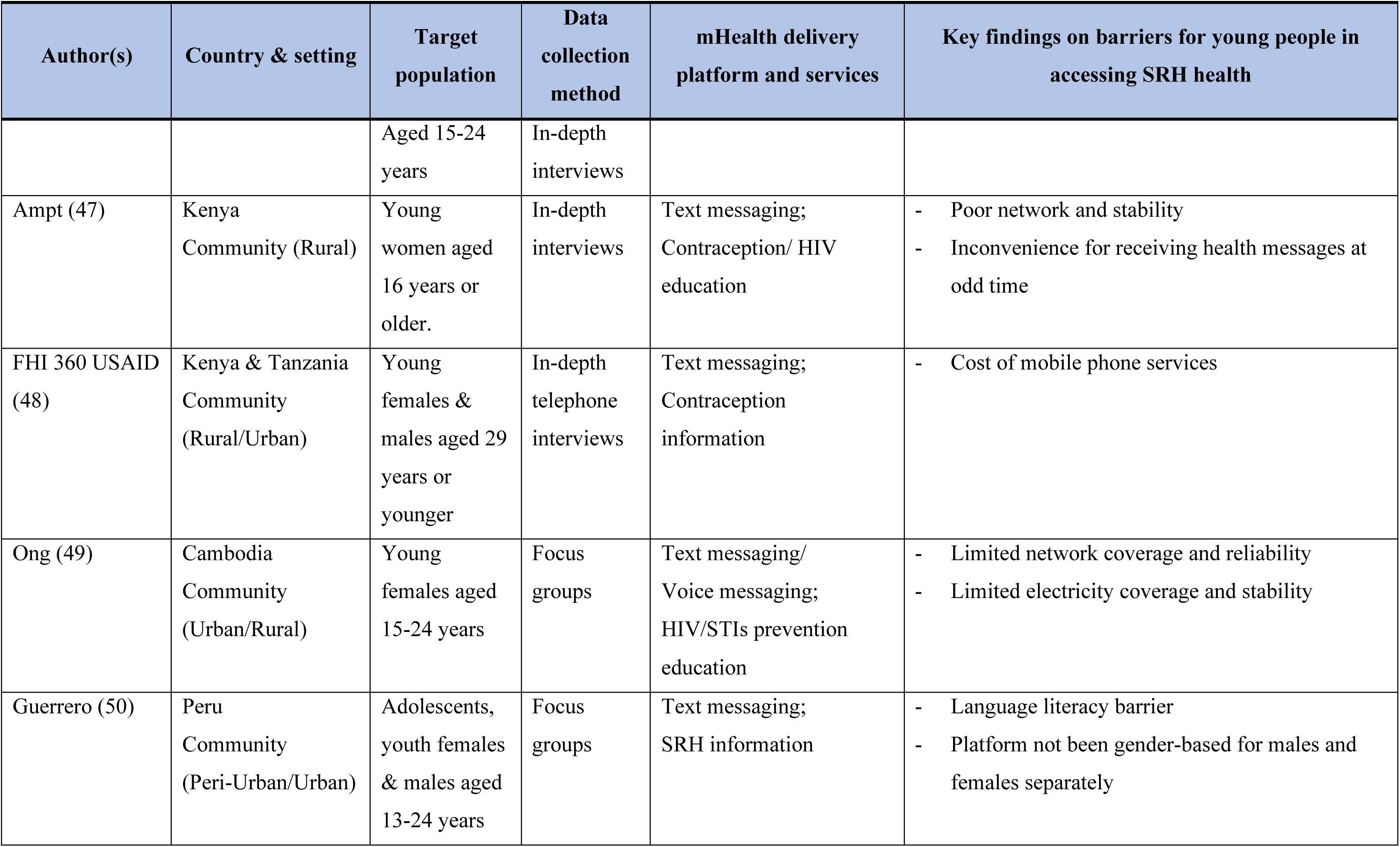
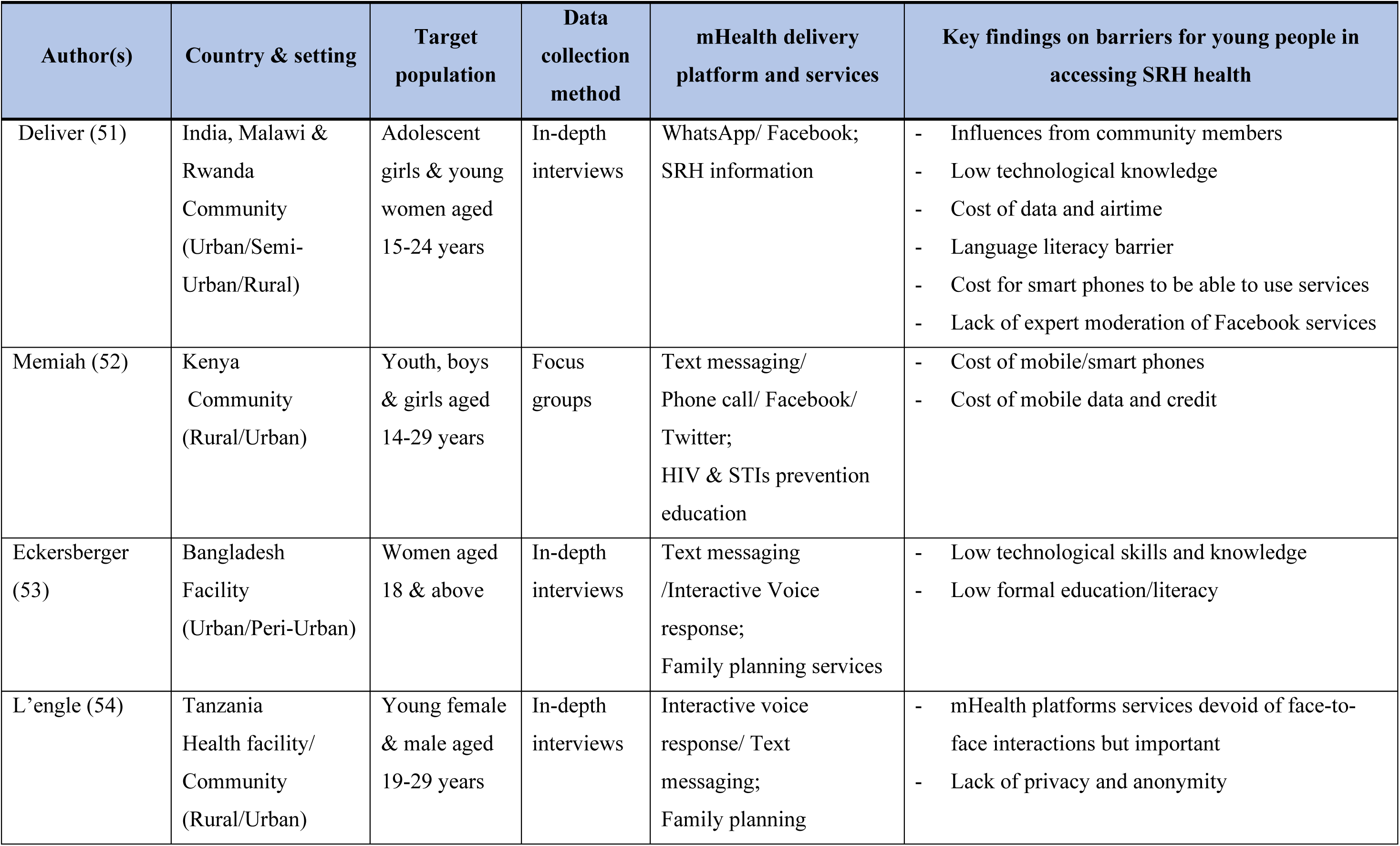
Summary of studies included in the systematic review.

The 26 studies included in this synthesis used a range of mobile phone platforms for providing access to SRH information and services among young people in rural settings in LMICs. Most of the studies used text-messaging platforms (11) with a combination of voice messaging, (2) interactive voice response, (2) phone calls, Facebook, WhatsApp and Twitter as noted in Table 5. The majority of studies included information about the barriers to mHealth SRH service access, these have also been included in summary form in Table 5.

### Confidence in the review findings

Confidence in the review findings across studies with explanation of the assessment are detailed in the CERQual evidence profile in Table 6. The study found high confidence in the findings for the text-messaging platforms (29, 30, 32, 34-38, 40, 42-50, 52-54). However, the confidence in the findings for social media platforms including WhatsApp (32, 42, 51) Facebook (32, 42, 51, 52) and Twitter (32, 52) was low. Generally, the study findings for confidence in the challenges and barriers for platform services was high.

**Table 6:** CERQual evidence profile.

| Summary of review findings | Studies contributing to review findings | CERQual assessment of confidence in the evidence | Explanation of CERQual assessment |
| --- | --- | --- | --- |
| **General experiences and perceptions for SRH mHealth platforms SRH information and services**<br>Participants had a wide range of experiences and views regarding interest, acceptability, and preferences for receiving mobile phone based SRH information and services. These were centred on comfort, convenience, privacy, confidentiality, anonymity, non-judgemental, ability for interaction, affordability and easier for accessing SRH messages compared to face-to-face services where communities are far away from health facilities reported across rural settings in LMICs. | (29-54) | High confidence | It is highly likely that the review finding is a reasonable representation of participants experiences for using mobile platforms SRH information and services. For the 26 studies, 3 had unclear ethical procedures, 3 had unclear reflexivity, 1 had unclear data analysis procedure and 3 with minor data issues. |
| **Experiences and perceptions for SRH mHealth platform SRH services** |  |  |  |
| **Text messaging**<br>Participants reported their preference for text messaging services due to the ability to store text messages for reference and share with friends and peers. Some participants also indicated being able to receive text messages under poor or weak network connectivity conditions as positive experiences for using the service. | (29, 30, 32, 34-38, 40, 42-50, 52-54) | High confidence | It is highly likely that the finding is a reasonable representation of participants experiences for text messaging. For the 21 studies, 2 had unclear ethical procedures, 1 an unclear data analysis procedure and 3 with minor data issues. |
| **Voice messaging**<br>Participant's ability to store voice messages and re-play and listen at their own convenience was expressed by some participants. Some participants | (30, 31, 33, 49, 53) | High confidence | It is highly likely that the finding is a reasonable representation of participants experiences for voice messaging. All the 5 studies had no or very minor concerns. |
| with low-literacy or no formal education expressed their preference for the service as it did not require writing and reading of text messages. |  |  |  |
| **Interactive voice response**<br>Most participants expressed positive views and preferences for interactive voice response due to the ability to interact or engage with health care providers or counsellors to discuss issues that were tailored to their needs. Young people with low education also indicated the ease of using the platform. The requirement for a basic mobile phone to use the services was reported as very advantageous to young poor rural people. | (29, 33, 40, 44, 45, 53, 54) | High confidence | It is highly likely that the finding is a reasonable representation of participants experiences for interactive voice response platforms. For all the 7 studies, one had minor data issues. |
| **Mobile phone call**<br>Participants reported positive experiences due to the ability to interact with health providers and to ask personal questions and get an instant response or feedback tailored to their needs. Low-literate young people liked the services for not requiring composing or reading text messages and not requiring smart or android phones to use the SRH intervention services. | (35, 38, 52) | Moderate confidence | It is possible that the finding is a reasonable representation of participants experiences for mobile phone call platform intervention SRH information and services. For all the 3 studies, ethical procedures was not clearly stated in one study. |
| **WhatsApp**<br>Participants reported positive experiences in terms of ability to interact with providers and friends and posting or sharing of questions on their | (32, 42, 51) | Low confidence | It is possible that the finding is a reasonable representation of participants experiences for WhatsApp platform SRH information and services due to limited data. For all the 3 |
| health issues on the platform for answers or responses. However, some expressed the services not been affordable due to the need for an android or smart phone to be able to use services as a challenge. |  |  | studies, ethical procedures in one of the studies were not clearly stated. |
| **Facebook**<br>Participants who used the services, a preference was related to the ability to interact with friends and peers and posting questions to receive answers or responses. Some participants reported that the requirement for an android and smart phone to be able to use the services was a barrier. Some had the perception that the services lacked expert moderation of the platform and information posted may not be authentic. | (32, 42, 51, 52) | Very low confidence | It is not clear whether the finding is a reasonable representation of participants experiences for Facebook platform SRH services due to thin data. However, for the 4 studies, the ethics procedures in one were unclear. |
| **Twitter**<br>Participants who used twitter platform services reported not being conversant with the platform services They also indicated that it familiar among the youth in their settings. Participants said they encountered some technical difficulties in manipulating the twitter handle. The need for an android or smart phone to be able to use services was also cited as a challenge. | (32, 52) | Very low confidence | It is not clear whether the finding is a reasonable representation of participants experiences for Twitter platform SRH services due to thin data. For the 2 studies, one had unclear ethics procedures. |
| **General barriers for use of mobile platforms SRH information and services by participants** |  |  |  |
| **Cost**<br>Participants cited cost for mobile phones and cost for mobile credit or airtime and data as a barrier to use of platforms services. In some instances, some had to depend on shared or borrowed phones to be able to access platforms services. Cost barrier was widely reported across several study settings. | (29-32, 34, 37, 38, 40-45, 48, 51, 52) | High confidence | It is highly likely that the finding is a reasonable representation of participants experiences of barriers in using SRH mobile phone platforms services. For all the 16 studies, one study had unclear ethical procedures, two with thin data and one with unclear reflexivity issues. |
| **Technological skills and language literacy**<br>Low technological knowledge, skills and language literacy were cited as barriers influencing fully use mobile phone platforms SRH services in across several rural settings | (29, 31-35, 37, 43, 44, 50, 51, 53, 55) | High confidence | It is highly likely that the finding is a reasonable representation of participants experiences of barriers in using SRH mobile phone platforms services. For all the 13 studies, two of the studies had unclear ethics procedures and unclear reflexivity. |
| **Electricity connectivity and stability**<br>Infrastructural barriers related to limited electricity connectivity and stability was cited a barrier affecting use of SRH mobile phone platforms services across several rural settings. This barrier was commonly reported across several rural settings. | (31, 32, 42, 44, 49) | High confidence | It is highly likely that the finding is a reasonable representation of participants experiences for mobile phone platforms SRH information and services. For all the 5 studies, one had unclear ethics procedures. |
| **Network connectivity and stability**<br>Infrastructural barriers related to limited and weak network connectivity and stability was cited a barrier affecting use of SRH mobile phone platforms services across rural settings. This barrier was commonly reported across several rural settings. | (29, 31, 32, 35, 40, 42, 44, 46, 47, 49) | High confidence | It is highly likely that the finding is a reasonable representation of participants experiences for SRH mHealth platforms SRH information and services. For all the 10 studies, 3 had unclear ethics procedure issues, one unclear reflexivity, and 2 with thin data. |
| **Social norms and beliefs**<br>Young people faced barriers as a result of community members influences. Parents and partners referred to women using SRH mHealth platforms services as been promiscuous. Some young people had the belief that receiving phone calls from unknown mobile phone contact numbers or persons could lead to death or the disappearance of the recipient. These restrictive beliefs and myths were reported across Tajikistan, Bolivia, Palestine, and Nigeria. | (31, 32, 36, 38, 44, 51) | Moderate confidence | It is highly likely that the finding is a reasonable representation of participants experiences for use of mobile phone platforms intervention SRH information and services. For all the 6 studies, 2 studies had issues with one was unclear reflexivity and the other on unclear ethical procedures. |

### Preference and acceptability for using mHealth SRH services

In this review, the studies used seven different mobile phone platforms to deliver a range of SRH information and services on contraception, family planning, prevention of STIs and HIV education across rural areas. Participants’ acceptance and preference levels for using mobile platform SRH intervention services were varied. Participants reported preferences for SRH text-messaging (29, 30, 32, 34-38, 40, 42-50, 52-54) services. Some of the reasons were associated with their ability to receive text messages even during poor or weak network connectivity conditions, the opportunity to store text messages for future reference and share the messages with friends and peers. Participants also reported a preference for using voice messaging services (30, 31, 33, 49, 53). Some of the reasons were due to their ability to be able to communicate with HCPs without writing, as well as the ability to store voice messages and re-play and listen to them when they needed. Recipients also reported that their ability to interact with HCPs or counsellors to gain quick responses to their SRH issues that were individually tailored to their needs also saw them favour voice messaging types of services over other methods.

The findings for the preference of phone call for SRH services (35, 38, 52) related to not requiring literacy and technical skills in terms for composing or reading text messages and the ability to use low cost basic mobile phones for the platform services.

### Barriers to using mHealth platforms SRH services

Barriers to using mobile platform services in rural settings centred on cost for mobile phones and their maintenance in terms of mobile credit or airtime and data, technological barriers related to lack of technological knowledge or skills needed to use platform services [160, 161, 165, 168, 173, 174, 178, 181, 186], limited infrastructure in terms of electricity and network connectivity and stability, and influence of social and cultural norms, influences from community members and beliefs and perceptions [161, 168, 181, 184, 186] as indicated in Table 2.6.

## Discussion

This review qualitatively explored the experiences and perceptions of young peoples’ use of mobile phone platforms for SRH information and services in rural settings in LMICs using the GRADE-CERQual approach. Evidence has shown that mHealth technology platforms have the potential to address numerous SRH information and services access barriers for young people in rural contexts in LMICs (16, 18, 56-59). The study findings reported preferences and barriers for use of mHealth-based platforms for SRH information and services among rural young people across rural LMICs.

Generally, cost in terms of mobile phone ownership and credit or data for their maintenance was reported as major barrier which influenced use of platforms SRH services among young people (29-32, 34, 37, 38, 40-45, 48, 51, 52). The cost factor led to the reliance on borrowed or shared phones by people who could not afford to be able to access SRH services. The use of borrowed and shared phones resulting in privacy and confidentiality challenges was also reported in this study [72, 105, 119, 120, 157, 162, 163, 168, 173] with high confidence in the study findings noted. Mobile phone sharing is a common phenomenon across rural settings in LMICs among poor young people and creates potential source of privacy and confidentiality breaches (60-63). This is a key challenge when discussing SRH information and services especially when concerning sensitive issues (44, 64). As such, young people are reluctant to use borrowed or shared phone for sensitive SRH issues (44). Ownership of a mobile phone guarantees independence and freedom for use for SRH information and services (44).

However, as rural young people are not economically independent, they may not be able to acquire and maintain mobile phones on their own for SRH information and services without financial interventions. Introduction of financial interventions is needed to make mobile platforms SRH services affordable for poor rural young people (64). Introducing financial subsidies for the cost of mobile phones and airtime for maintenance through flexible terms of payment could help overcome the cost barrier and sustain use of services among young people in rural settings in LMICs (65).

The review finding reported limited and poor network (29, 31, 32, 35, 40, 42, 44, 46, 47, 49) and electricity connectivity and coverage (31, 32, 42, 44, 49) as a barriers to use of mobile platforms SRH information and services among young people across the study settings. This is consistent with other studies which have identified limited coverage and poor network and power connectivity as a challenge in rural LMICs to the access of SRH information and services with young people struggling to identify spots in the communities with network connectivity and stability to be able to use mobile phones (64, 66-69). In addition, charging phones often requires travel to communities with power and sometimes paying a fee to charge mobile phones batteries. Given that the use of mHealth applications in rural settings requires a reliable, effective, and sustained network and electricity connectivity (17, 70), exploring alternative power sources such as solar panels and power banks have been recommended for rural poor-resourced settings in LMICs with limited technological infrastructure (17, 65).

High confidence was found in studies reporting on limited technological skills and language literacy (29, 31-35, 37, 43, 44, 50, 51, 53, 55) as barriers to mHealth for SRH particularly in terms of limited knowledge regarding the use of these platforms. Despite the perceived benefits and opportunities of mobile phone technology for SRH information services among rural populations (especially young people in rural communities in LMICs), these services are less utilisable by low literate and illiterate young people in rural contexts that would otherwise have benefited most from mHealth technologies SRH initiatives (71).

Moderate confidence was shown in the findings that identified the restriction of young people’s mobile phone use for SRH services by community members such as partners and parents (31, 32, 36, 38, 44, 51) as a barrier to use. This finding is consistent with other studies in rural LMICs where young women reported that their sexual partners associated phone use for SRH information with infidelity (62, 63, 66). Despite the untapped potential of mobile phones as tools for improving access to SRH information and services among young people in rural LMICs, the use of mobile phones frequently raise a topic of conflict between sexual partners(66) and could influence the decision of young people to use innovative mHealth technology for SRH information and services. Other perceived factors which influenced young people’s phone use for SRH services emanated from social beliefs and myths related to phone calls received from unknown people that could result in death or disappearance of the recipient. This was most often reported across rural settings in Tajikistan, Bolivia Palestine, and Nigeria. Other studies have also linked social beliefs and myths for receiving calls from anonymous people resulting in death caused by “evil spirits” (62, 63). These social beliefs need to be addressed to avoid misinforming young people for using the mobile phone technology for SRH information and services in rural context of LMICs. Public education is critical to educate rural populations about mHealth SRH initiatives to address knowledge gaps in relation to myths and perceptions that affect use of mobile phone platforms for SRH information and services among young people in rural contexts in LMICs (72, 73).

The study also reported positive findings regarding the acceptance and preferences of mHealth platforms SRH information and services in relation to youth-friendly platforms services (58, 74, 75) associated with comfort, convenience, privacy and confidentiality, compared to in-person services (29-54). The ability to engage young people with wide range of SRH information and services across wide geographical rural areas were reported as some of the benefits of mHealth platforms for SRH services (29-54). Several studies have come out with similar findings among young people in rural contexts in LMICs (6, 76-79). With regard to platforms such as text messaging, voice messaging, integrated voice recognition (IVR) and phone calls, some of the positive findings in terms of their preference for SRH information and services was due to their familiarity and the ability to use low cost non android phones for the services, compared to WhatsApp, Facebook, and Twitter platforms. The preference for text-messaging services was associated with the ability to receive and read text messages under poor or weak network conditions and the opportunity to store messages for reference and share with friends and peers (29, 30, 32, 34-38, 40, 42-50, 52-54). These findings are also confirmed by studies in rural settings in LMICs among young people (6, 50, 64, 80-83). Text-messaging platforms continue to be the most frequently used and preferred mobile phone communication platform among populations in rural settings in LMICs (44). However, text messaging requires basic literacy for writing and reading to be able to use services, hence young people’s preference for phone calls (35, 38, 52) and voice messaging (30, 31, 33, 49, 53) and IVR (29,

33, 40, 44, 45, 53, 54) services. Preference for voice messaging, phone call, and IVR platforms was also due to illiterate and low literate young people to use the platform services since they did not require writing and reading of messages (30, 31, 33, 49, 53) and the to the ability to interact or engage with HCPs or counsellors to discuss their SRH issues by receiving quick response tailored to their needs (29, 33, 40, 44, 45, 53, 54). The findings indicated that platforms such as WhatsApp (32, 42, 51), Facebook (32, 42, 51, 52) and Twitter (32, 52) were less preferred and used by young people for SRH information and services due to their unfamiliarity and the requirement for a smartphone and its associated high cost as well as low knowlededge and skills (29-32, 34, 37, 38, 40-45, 48, 51, 52).

While basic or simple low cost mobile phone ownership levels are common among rural young people in LMICs and could be used for mHealth SRH initiatives, the same is not true for smartphones (84, 85). Whiles mobile phone-based platforms are becoming increasingly popular among young people for SRH services in rural contexts in LMICs, platforms such as WhatsApp, Facebook and Twitter are less used and preferred by young people (42, 51, 52). Few young people had used these platforms due to the requirement for smart phones and their associated high cost and lack of technological skills (16, 86, 87). Some participants also felt that these platforms were not familiar among them in their settings and hence their low usage. Evidence has shown that there is a dearth of qualitative evidence on the effect of innovative social media platforms such WhatsApp, Facebook, and Twitter for SRH information and services among young people in rural LMICs (42, 51, 52). This warrants further research on these social media platforms to help maximise their impact for use for SRH information and services among young people in rural LMICs settings.

We acknowledge that this review has some limitations. Firstly, we restricted our search to those studies published in the English language between 2000-2020. This could have limited the numbers of articles restricted from the search. We could have missed relevant articles outside of our study period and languages other than English. Hence, we could have missed studies relevant to mHealth platforms for SRH services for young people. A strength of this study is that it was conducted using a comprehensive search strategy and a robust quality appraisal process and rigorous systematic review methodology (25-28).

## Conclusion

The findings of this synthesis have demonstrated the perceived potential of mHealth platforms for improving access to conventional SRH information and services among young people in rural settings in LMICs. While mHealth platforms have the potential to address access limitations to SRH information and services among rural young people, the barriers need to be addressed. These findings provide relevant recommendations for policy makers and implementers of SRH programs to improve services for integration and scale-up in the rural health system contexts in LMICs.

## Authors’ contributions

ASL, DL and MLH conceived and designed the study. ASL, DL and MLH conducted the data search. ASL and NK conducted the title and abstract screening, full text screening and data extraction. ASL and NK analysed the data and conducted the quality assessments. Data screening was independently conducted by two reviewers (ASL and NK) based on title and abstract according to the inclusion criteria. Full text articles were retrieved, and the two reviewers (ASL and NK) independently assessed the relevancy of the papers. Screened articles were read in full. Discrepancies were resolved by discussion. ASL wrote the first draft of the manuscript. ASL, NK, MLH and DL assessed the confidence of the review findings. MLH, DL and NK contributed to the interpretation of the data and review of the manuscript for intellectual content. All the authors have read and approved the manuscript for submission.

## Data Availability

Data is available from the corresponding author upon a reasonable request.

## Acknowledgements

This research was supported by an Australian Government Research Training Program (RTP) Scholarship for my PhD program. We would like to acknowledge Jessica Birchall, senior librarian at the University of Newcastle for her guidance in developing the search terms for this review. Dr. Melissa Harris is supported by an ARC Discovery Early Career Researcher Award ((DE190101134).

## Availability of data and materials

The datasets used and/or analysed during the current study are available from the corresponding author on reasonable request.

## Appendix

**Table1:** Search strategies tailored to the other databases.

| Cochrane Library |  |
| --- | --- |
| 1 | Trials matching young adult* OR youth* OR adolescent* OR young people* OR youth population* OR young wom?n* OR young girl* OR young boy* OR young m?n* OR young women* emerging adult* OR adolescent girl* OR adolescent boy* OR adolescen |
| 2 | In 'Title Abstract Keyword' AND reproductive health* OR sexual health* OR HIV* OR contraception* OR contraceptives* OR modern contraception* OR contracept service* OR contracept educat* OR contracept counsell |
| 3 | In 'Title Abstract Keyword' OR Healthcare providers* OR Healthcare professionals* OR health provider* OR health counsellor* OR health educator |
| 4 | In 'Title Abstract Keyword' AND mobile health* OR mHealth* OR mobile phone health technology* OR mobile phone health* OR digital mobile health* OR digital mobile phone health |
| 5 | In 'Title Abstract Keyword' AND low-income countries* OR low-and-middle-income nation* OR low to middle income countries* OR middle-income countr* OR low resource countries |
| 6 | In 'Title Abstract Keyword' - with Cochrane Library publication date Between Jan 2000 and Dec 2020, in Trials with 'Public Health' in Cochrane Groups (word variations have been searched) |
| Scopus |  |
| 1 | (TITLE-ABS-KEY (low-income AND countries* OR low-and-middle income AND nation* OR low AND toAND middle AND income AND countries* OR middle-income AND countr* OR low AND resource AND countries) |
| 2 | TITLE-ABS KEY (mobile AND health* OR mhealth* OR mobile AND phone AND health AND technology* OR mobile AND phone AND health* OR digital AND mobile AND health* OR digital AND mobile AND phone AND health) OR |
| 3 | TITLE-ABS-KEY (healthcare AND providers* OR healthcare AND professionals* OR health AND provider* OR health AND counsellor* OR health AND educator ) OR |
| 4 | TITLE-ABS-KEY (young AND adult* OR youth* OR adolescent* OR young AND people* OR youth AND population* OR young AND wom?n* OR young AND girl* OR young AND boy* OR young AND m?n* OR young AND women* AND emerging AND adult* OR adolescent AND girl* OR adolescent AND boy* OR adolescen) |
| 5 | TITLE-ABS-KEY (reproductive AND health* OR sexual AND health* OR HIV* OR contraception* OR contraceptives* OR modern AND contraception* OR contracept AND service* OR contracept AND educat* OR contracept AND counsell) |
| 6 | AND PUBYEAR > 1999 AND PUBYEAR < 2021. |
| **CINAHL** |  |
| S4 | (reproductive health* AND sexual health* AND HIV* AND contraception* AND contraceptives* AND modern contraception* AND contracept service* AND contracept educat* AND contracept counsell* AND young adult* AND youth* AND adolescent* AND young people* AND youth population* AND young wom?n* AND young girl* AND young boy* AND young m?n* AND young women* AND adolescent girl* AND adolescent boy* AND adolescen) OR (healthcare providers* AND healthcare professionals* AND health provider* AND health professional |
| S3 | (reproductive health* OR sexual health* OR HIV* OR contraception* OR contraceptives* OR modern contraception* OR contracept service* OR contracept educat* OR contracept counsel* OR young adult* OR youth* OR adolescent* OR young people* OR youth population* OR young wom?n* OR young girl* OR young boy* OR young m?n* OR young women* emerging adult* OR adolescent girl* OR adolescent boy* OR adolescen |
| S2 | (reproductive health* OR sexual health* OR HIV* OR contraception* OR contraceptives* OR modern contraception* OR contracept service* OR contracept educat* OR contracept counsel* OR young adult* OR youth* OR adolescent* OR young people* OR youth population* OR young wom?n* OR young girl* OR young boy* OR young m?n* OR young women* emerging adult* OR adolescent girl* OR adolescent boy* OR adolescen ) AND ( mobile health* OR mHealth* OR mobile phone health technology* OR mobile phone health |
| S1 | (reproductive health* OR sexual health* OR HIV* OR contraception* OR contraceptives* OR modern contraception* OR contracept service* OR contracept educat* OR contracept counsel* OR young adult* OR youth* OR adolescent* OR young people* OR youth population* OR young wom?n* OR young girl* OR young boy* OR young m?n* OR young women* emerging adult* OR adolescent girl* OR adolescent boy* OR adolescen ) AND ( mobile health* OR mHealth* OR mobile phone health technology* OR mobile phone health* |
|  | limiters - full text; references available; published date: 20000101-20201231; peer reviewed; clinical queries: qualitative - best balance; human; journal subset: public health; geographic subset: Africa; language: English; special interest: men's health, public health, women's health |
| **PsychoINFO** |  |
| 1 | (young adult* or youth* or adolescent* or young people* or youth population* or young wom?n* or young girl* or young boy* or young m?n* or young women* emerging adult* or adolescent girl* or adolescent boy* or adolescen).mp. [mp=title, abstract, heading word, table of contents, key concepts, original title, tests & measures, mesh] |
| 2 | (reproductive health* or sexual health* or HIV* or contraception* or contraceptives* or modern contraception* or contracept service* or contracept educat* or contracept counsell).mp. [mp=title, abstract, heading word, table o'f contents, key concepts, original title, tests & measures, mesh] |
| 3 | (healthcare providers* or healthcare professionals* or health provider* or health counsellor* or health educator).mp. [mp=title, abstract, heading word, table of contents, key concepts, original title, tests & measures, mesh] |
| 4 | (mobile health* or mHealth* or mobile phone health technology* or mobile phone health* or digital mobile health* or digital mobile phone health).mp. [mp=title, abstract, heading word, table of contents, key concepts, original title, tests & measures, mesh] |
| 5 | (low-income countries* or low-and-middle-income nation* or low to middle income countries* or middle-income countr* or low resource countries).mp. [mp=title, abstract, heading word, table of contents, key concepts, original title, tests & measures, mesh] |
| 6 | limit to (full text and human and english language and yr="2000 - 2020") |

**Table2:**
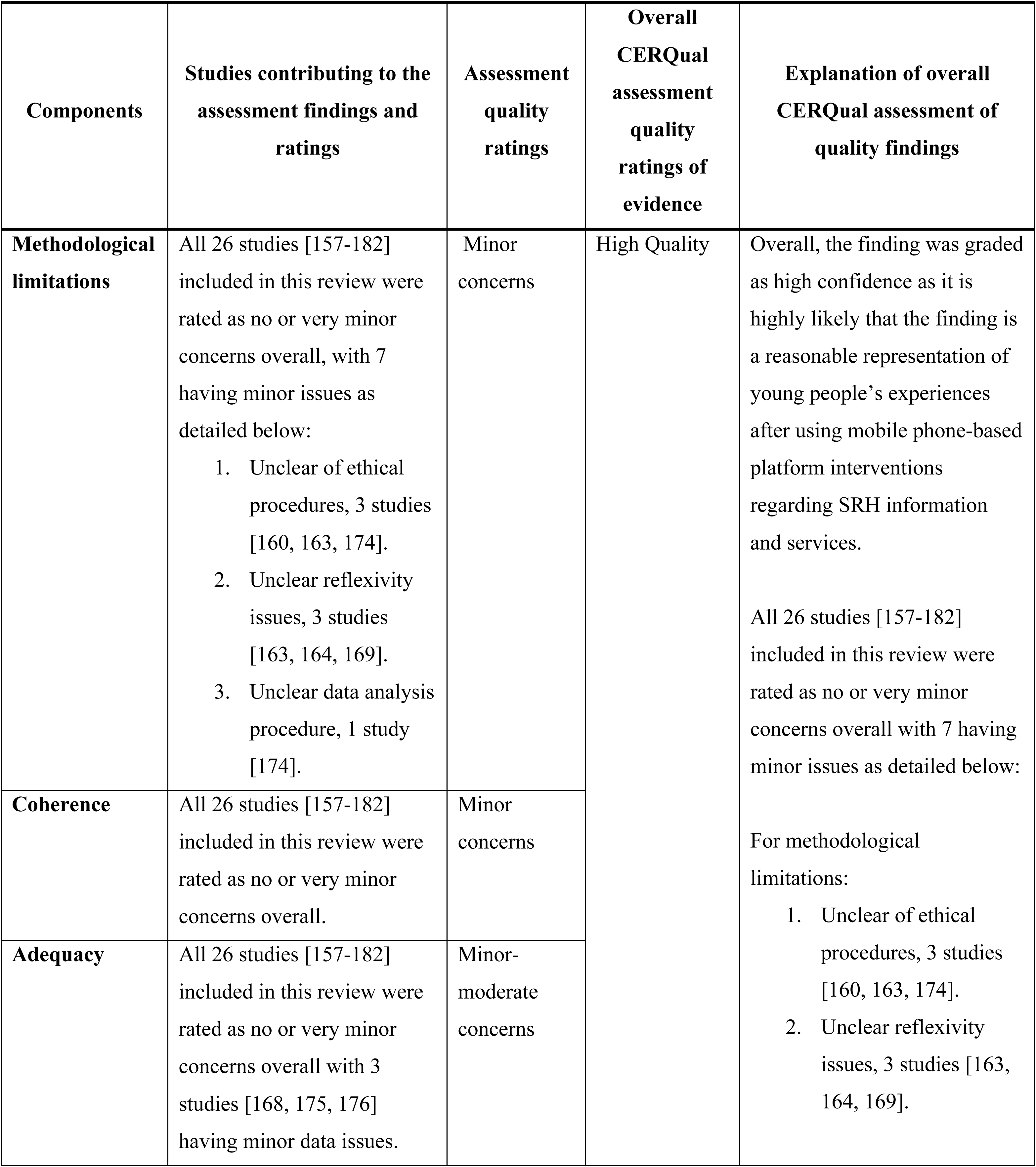
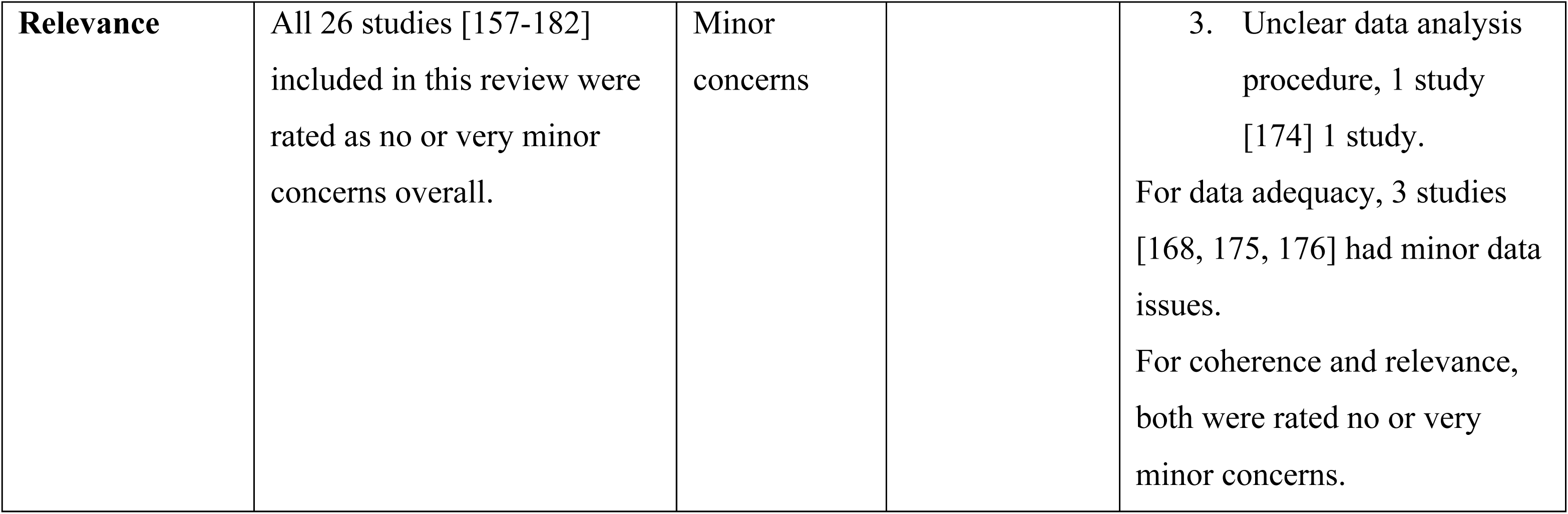
Overall summary of CERQual assessment of quality.

**Table3:**
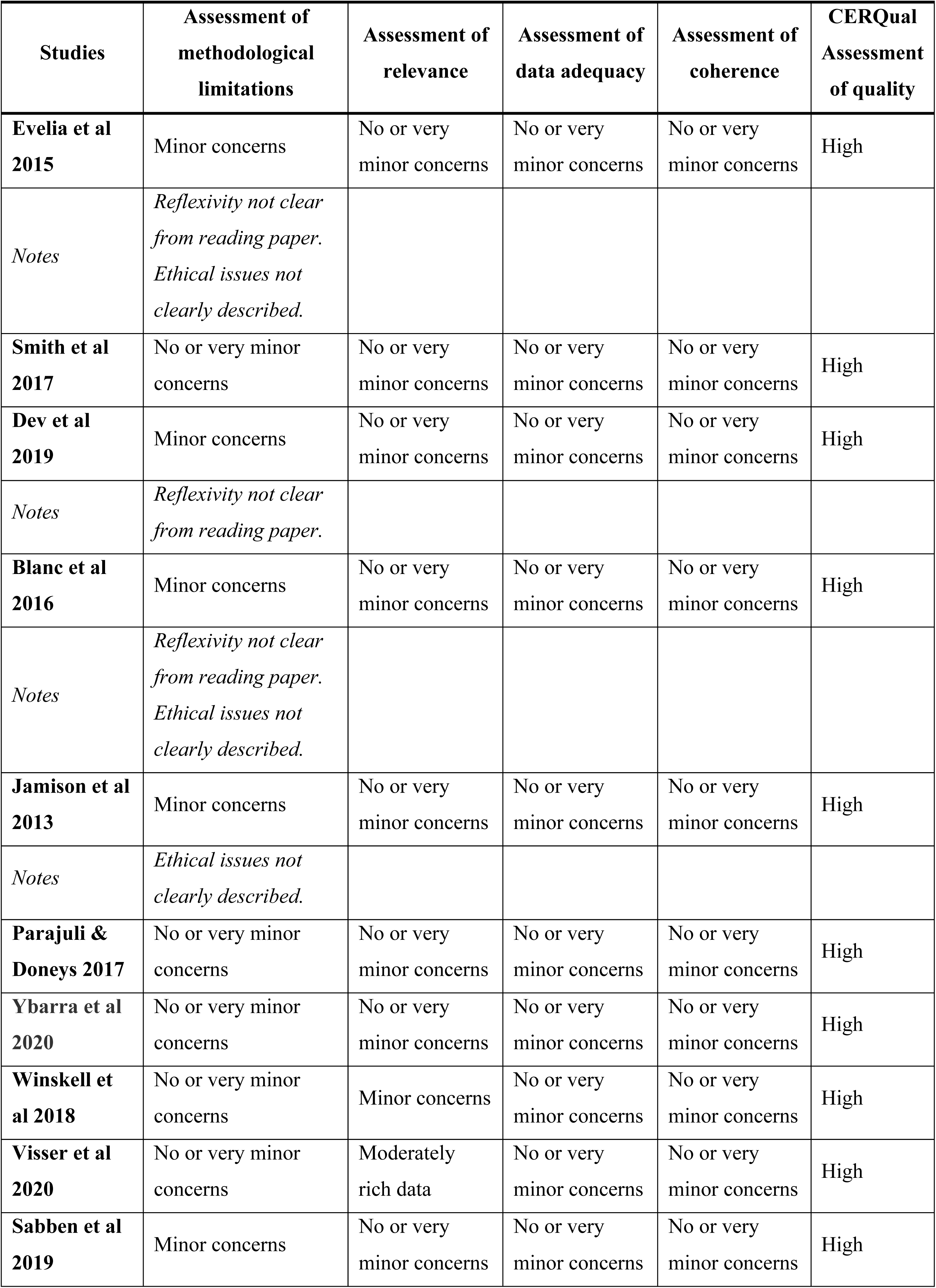
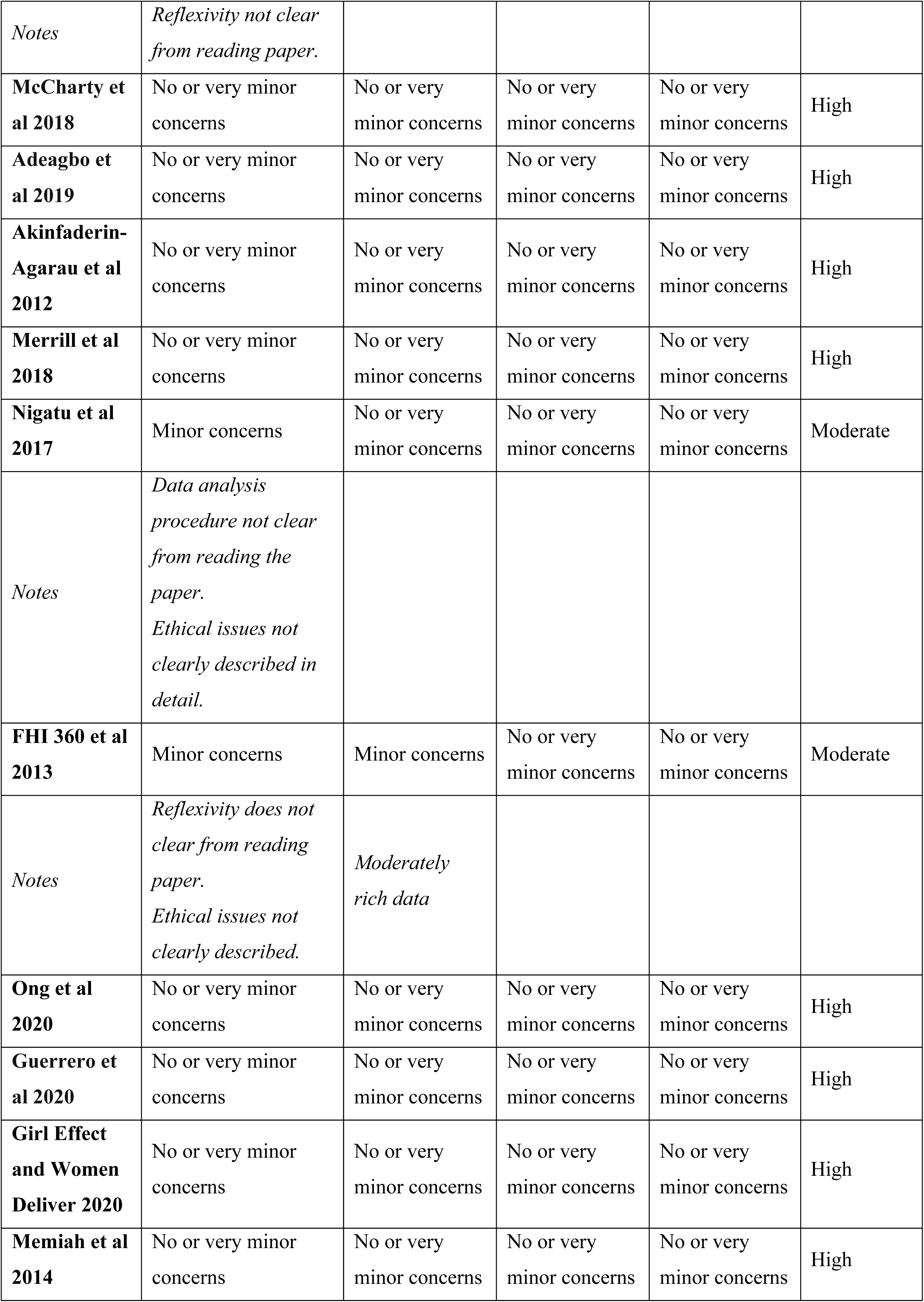
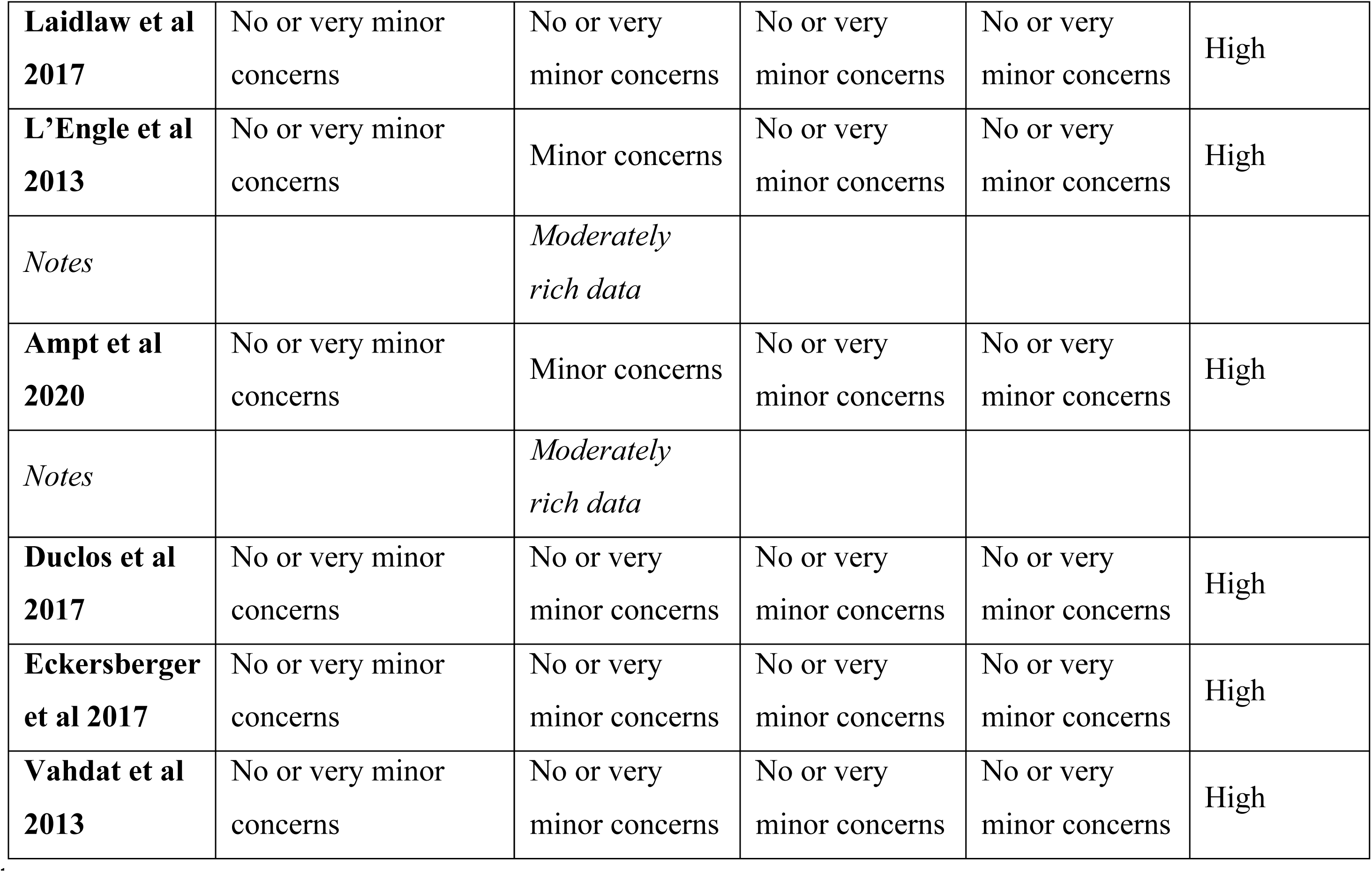
Assessment of studies under each CERQual component.

